# Evaluating the Generalizability of EEG-Based AI Models in Alzheimer’s and Dementia Diagnosis

**DOI:** 10.1101/2025.09.10.25334048

**Authors:** Rajkumar Saini, Foteini Liwicki, Sumit Rakesh, Hamam Mokayed, Sarthak Acharya, Daljeet Singh, Vibha Gupta

## Abstract

**INTRODUCTION:** We thoroughly investigated the generalizability of deep learning models trained on electroencephalography (EEG) data to detect Alzheimer’s disease and dementia at the individual subject level. Although average model performance appears strong, it may obscure large inter-individual variability, raising concerns for clinical deployment.

**METHODS:** We trained a Hopfield-enhanced deep neural network on a publicly available EEG dataset consisting of 88 participants, including individuals diagnosed with Alzheimer’s disease (AD), frontotemporal dementia (FTD), and cognitively normal controls (CN). Resting-state EEG recordings were segmented and used to train the model in a leave-onesubject-out (LOSO) cross-validation setup across multiple detection tasks: AD vs. CN, AD vs. FTD, FTD vs. CN, and AD vs. FTD vs. CN.

**RESULTS:** While the model demonstrated high average performance (e.g., up to 83% accuracy), subject-level results revealed inconsistencies. Some individuals achieved perfect prediction even at the first training epoch, suggesting spurious memorization, while others predicted falsely throughout, with performance below chance. These patterns persisted despite consistent training conditions and no data leakage.

**DISCUSSION:** Our findings highlight that strong group-level performance may be misleading in clinical settings, where decisions are made at the individual level. The models should be generalizable across individuals and be evaluated per individual before being considered for diagnostic use. Hopfield networks show promise in capturing patterns in EEG data, but patient-level validation and transparent reporting are essential to avoid premature clinical translation.

**Highlights:**

- Deep learning models trained on EEG can achieve high average performance in detecting Alzheimer’s and dementia disease.
- The subject-level evaluation revealed significant variability, including below-chance performance for some individuals despite overall strong results.
- Group-level metrics alone may be misleading; Generalizable models and individual-level validation are critical before the clinical adoption of EEG-based AI models.

## 1 | INTRODUCTION

Alzheimer’s disease (AD) and frontotemporal dementia (FTD) are progressive neurodegenerative disorders that pose growing public health concerns around the world [1]. Accurate early-stage diagnosis remains a clinical priority, particularly given the limitations of current biomarkers, which may require invasive procedures or be cost-prohibitive for routine use [2, 3]. In this context, electroencephalography (EEG) has gained renewed attention due to its non-invasive, inexpensive, and temporally precise nature, making it a promising tool for scalable Alzheimer’s / dementia detection and monitoring [4].

Recent advances in deep learning have further propelled interest in EEG-based biomarkers for the detection of Alzheimer’s and dementia. Deep neural networks can learn complex spatiotemporal patterns in EEG signals without hand-crafted features, often exceeding traditional machine learning in performance. Multiple studies have demon-strated the potential of convolutional neural networks (CNNs), recurrent architectures, and hybrid models to distinguish AD or FTD from cognitively normal individuals (CN) [5, 6, 7]. For instance, Miltiadous et al. [5] proposed DICE-Net, a dual-input convolution-transformer architecture that combines relative band power and spectral coherence features for Alzheimer’s detection from EEG signals. Their model achieved superior accuracy (83.28%) compared to conventional machine learning and CNN-based methods, validated with Leave-One-Subject-Out (LOSO) strategy on a publicly available dataset.

Stefanou et al. [6] proposed a CNN using FFT-based spectrogram representations and achieved over 79% accuracy on AD/CN prediction in a subject-wise evaluation framework. Similarly, Zhang et al. [7] applied deep learning to EEG for AD diagnosis and highlighted the challenge of inter-patient variability, despite promising average performance.

However, a critical issue persists across much of the literature: the tendency to report group-level performance metrics, such as mean accuracy, without sufficiently examining individual subject outcomes. In real-world clinical settings, models must generalize to new patients, making subject-level reliability essential. When models are evaluated via cross-validation schemes like k-fold or leave-group-out, average performance may appear robust even if the model fails catastrophically for certain individuals. AlSharabi et al. [8] acknowledged this challenge in a clinical decision support system for AD, noting a wide variation in performance under LOSO cross-validation despite a strong average accuracy.

In our study, we aimed to explore this issue systematically. We trained a Hopfield-enhanced deep neural network on a publicly available EEG dataset of 88 subjects that includes AD, FTD, and CN participants. Hopfield networks, which offer associative memory mechanisms and pattern stability properties, have shown potential for modeling temporal EEG dynamics in our study. Using Leave-One-Subject-Out (LOSO) cross-validation, a gold standard for evaluating subject generalizability, we conducted four prediction tasks: AD vs. CN, AD vs. FTD, FTD vs. CN, and AD vs. FTD vs. CN.

Our findings are both comparable with existing works and cautionary. While overall accuracy reached up to 83%, the subject-level results revealed striking inconsistencies. Certain individuals achieved 100% accuracy even at the first epoch of training, suggesting potential overfitting, data imbalance, or spurious correlations. Conversely, other subjects consistently yielded accuracies below chance, even after prolonged training. These extremes occurred under controlled conditions and identical hyperparameters, with no evidence of data leakage.

This study highlights a critical gap in the current EEG-AI research: the overreliance on average performance metrics can obscure meaningful subject-level variability. For models intended for clinical use, such omissions carry real-world risks, where a seemingly robust algorithm may fail unpredictably for individual patients. By systematically examining per-subject performance trajectories, we demonstrate (from the structured experimental logs) the value of granular validation in revealing hidden patterns of false prediction or spurious generalization. These insights advocate for more transparent and individualized evaluation frameworks as a prerequisite for the safe and ethical translation of deep learning models into Alzheimer’s and dementia care.

This study contributes the following:

- Hopfield Network-based deep learning models are suitable to model Alzheimer’s detection.
- Subject-level evaluation using LOSO reveals variability hidden by average performance metrics.
- High average accuracy can coexist with complete failures for certain individuals, posing a risk for clinical deployment.
- Reporting per-subject performance is essential for reliable clinical translation of EEG-based AI models.
- Threshold-based epoch ranking ensures selection of well-trained and more-generalized models, accounting for the fact that the optimal epoch can vary across individual subjects in LOSO evaluation.
- Experiments on a public dataset along with the provided code and structured per-subject training logs, enables full reproducibility and transparency of the experiments.

## 2 | MATERIAL

### 2.1 | Dataset Description

Resting-state EEG recordings were obtained at the 2nd Department of Neurology, AHEPA General Hospital of Thessaloniki, using a Nihon Kohden EEG-2100 system, by the data curators. Nineteen scalp electrodes were placed according to the international 10-20 system, with mastoid references (A1, A2) used for impedance monitoring. Data acquisition followed a standardized clinical protocol with subjects seated, eyes closed, and electrode impedance maintained below 5 *k* Ω.

Signals were sampled at 500 Hz with a sensitivity of 10 *µV* /*mm*. The referential montage, using Cz as the common reference, was made publicly available. Recording parameters included a time constant of 0.3 seconds and a hardware-defined frequency range of 0.5-70 Hz. Average recording durations were 13.5 minutes for AD (range: 5.1-21.3 min), 12 minutes for FTD (7.9-16.9 min), and 13.8 minutes for CN (12.5-16.5 min), yielding a total of approximately 1,164 minutes of EEG data across all subjects.

Preprocessing followed a standardized pipeline. EEG signals were first band-pass filtered between 0.5 and 45 Hz (Butterworth filter) and re-referenced to A1-A2. Artifact correction was performed using the Artifact Subspace Reconstruction (ASR) method in EEGLab with a conservative 17 standard deviation threshold over 0.5-second windows. Independent Component Analysis (ICA) was then applied, and components classified as ocular or jaw artifacts were automatically rejected using the ICLabel plugin. The resulting signals were stored in BIDS-compatible .set format, organized by subject.

### 2.2 | Participants

The original study included 88 adult participants, categorized into three diagnostic groups: 36 with Alzheimer’s disease (AD), 23 with frontotemporal dementia (FTD), and 29 cognitively normal controls (CN). Diagnoses were established through clinical and neuropsychological assessments, consistent with standardized diagnostic criteria for dementia syndromes.

Cognitive status was evaluated using the Mini-Mental State Examination (MMSE), a validated measure of global cognitive function. The mean MMSE scores reflected the expected cognitive gradient: 17.75 (SD = 4.5) for AD, 22.17 (SD = 8.22) for FTD, and 30 for CN. The duration of the disease among dementia participants had a median of 25 months (interquartile range: 24 to 28.5 months). No comorbid neurodegenerative diagnoses were reported in the AD group. The average age at the time of EEG recording was 66.4 years (SD = 7.9) for AD, 63.6 years (SD = 8.2) for FTD, and 67.9 years (SD = 5.4) for participants in the CN.

### 2.3 | Ethics

This study used publicly available, de-identified EEG data from the OpenNeuro repository (dataset ID: ds004504). As the dataset is fully anonymized and open-access, no additional ethical approval or informed consent was required for this secondary analysis. All procedures adhered to the data use terms specified by the original dataset authors and the OpenNeuro platform.

## 3 | METHOD

In this section, we describe the deep learning (DL) pipeline developed for the experiments, including the model architecture, layer configurations, input and output dimensions, and the objective loss function. We also provide details on the computational resources used for model training and evaluation.

### 3.1 | Deep Learning Pipeline

#### 3.1.1 | Hopfield-Enhanced DL Architecture

To model EEG-based Alzheimer’s and dementia detection from EEG signals, we designed a custom deep neural network architecture termed `HopfieldEEGNet5`, combining convolutional feature extraction with Hopfield-based associative memory layers. The model was implemented in PyTorch and trained using GPU acceleration.

#### Input Representation

The model accepts EEG segments as input tensors of shape [*B*, 1, *C*, *T*], where *B* is the batch size, *C* = 19 is the number of EEG channels, and *T* = 15000 corresponds to the temporal length (30 second) of each segment.

##### Convolutional Backbone

The first module comprises a stack of four 2D convolutional layers:

- Layer 1: Conv2D (1→8) with kernel size (19, 11), stride (1, 2), and padding (0, 5), followed by BatchNorm, Sigmoid activation, and Dropout (*p* = 0.3).
- Layer 2-4: Subsequent Conv2D layers with increasing channel depth (8→16→32→64), each followed by BatchNorm and Sigmoid activation, maintaining kernel size along the time axis to progressively reduce temporal resolution.
- Final convolutional output is passed through an AdaptiveMaxPool2D layer to standardize output shape to [*B*, 1, 64].

Hopfield Layer

The flattened convolutional output is enriched with learnable positional encodings and normalized using LayerNorm. This is followed by a Hopfield layer configured with:

- `input_size = 64,`
- `stored_pattern_size = 64,`
- `pattern_projection_size = 64,`
- `update_steps_max = 3.`

This module supports pattern stabilization and temporal memory integration across the feature sequence.

**FIGURE 1.**
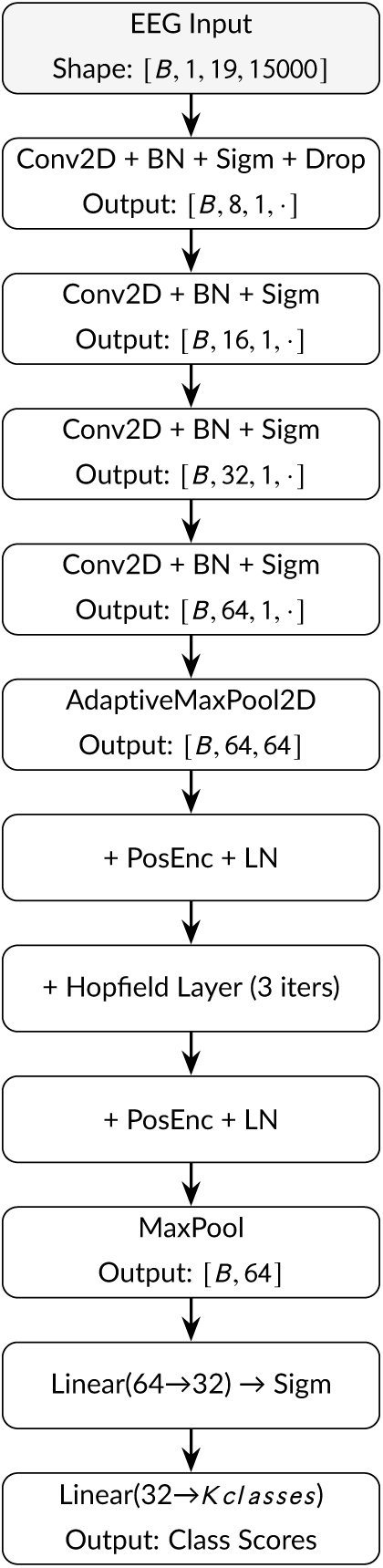
Architecture of the HopfieldEEGNet5 model.

##### Classification Module

The Hopfield-enhanced representation undergoes LayerNorm and max pooling across the sequence dimension. The resulting vector is passed through a two-layer fully connected classifier (Linear: 64→32→*n_k_*) with intermediate Sigmoid activation. The output dimension reflects the specific prediction task (binary or multiclass).

#### 3.1.2 | Performance Metrics

Model evaluation was performed using standard performance metrics as implemented in the `scikit-learn` library:

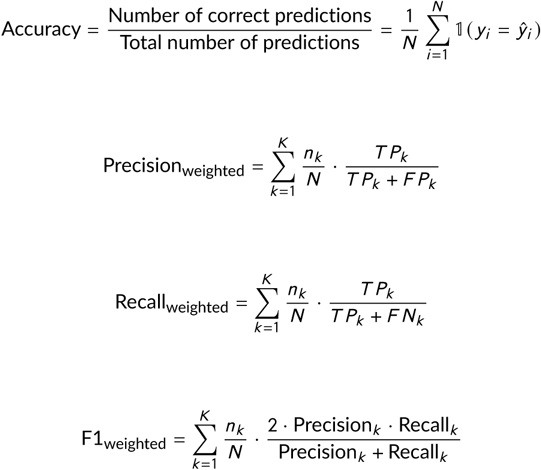

In the above:

- *N* is the total number of samples,
- *K* is the number of classes,
- *n_k_* is the number of samples in class *k*,
- *T P_k_*, *F P_k_*, *F N_k_* are the true positives, false positives, and false negatives for class *k*.

#### 3.1.3 | Training Configuration and Computational Resources

Model training was conducted using the Adam optimizer with a learning rate of 1×10^−3^ and a categorical cross-entropy loss function. The network was trained for a fixed number of epochs=51 across all cross-validation folds, with no early stopping or hyperparameter tuning. Each fold corresponded to a Leave-One-Subject-Out (LOSO) setting, in which the model was trained on *n* − 1 subjects and evaluated on the held-out individual. This ensured strict subject-level independence during evaluation.

We used a `batch size` of 16 and set the data shuffling seed to 42 to ensure reproducibility. Data were loaded using subject-stratified dataloaders, and the integrity of the LOSO setup was programmatically verified before each fold to prevent data leakage. During training, each mini-batch was passed through the model in training mode, and gradients were updated using backpropagation. The model’s performance was logged at every epoch, including training and test accuracy, precision, recall, F1-score, and confusion matrix. To monitor generalization, the model state was checkpointed only if it showed both improved test accuracy and reduced training loss compared to previous epochs.

Training was performed on an NVIDIA A100 GPU with 40 GB of memory. The logs were maintained to simultaneously write outputs to the terminal and timestamped log files, ensuring traceability across experiments.

## 4 | RESULTS

In this section, we show the results of the study for binary and multiclass settings: AD/CN, FTD/CN, AD/FTD, and AD/FTD/CN. For each subject under LOSO validation, we trained the AI model for 51 epochs. It is to be noted that the model’s *loss* should ideally decrease as training progresses, and other performance metrics *accuracy, precision, recall, F1* are expected to increase. Each of these models was ranked (highest rank=1) considering *training loss, and training & testing F1 scores*. So, that higher training loss lowers the rank even if the model has relatively higher F1 scores.

### 4.1 | AD/CN

Table 1 shows a fine-grained analysis of AD/CN results under LOSO validation; rather than just reporting an overall average performance, Table 1 also shows which subjects fall in different accuracy bins.

**TABLE 1.**
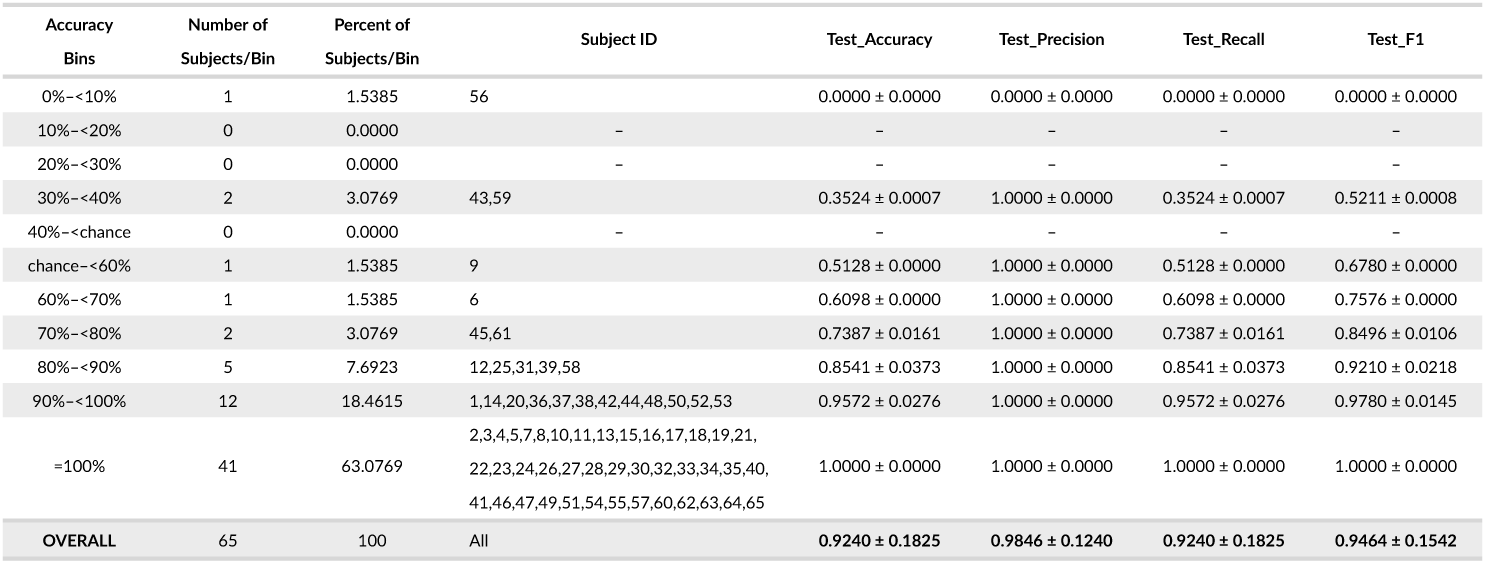
AD/CN-Binning of top-1 epoch for each subject-Further investigating.

For each subject (*n* = 65) we selected the single checkpoint with the highest test F1 and binned subjects by their Top-1 test accuracy (Table 1). The distribution is strongly right-skewed: *41/65 (63.1%)* subjects achieved *perfect* performance (Accuracy/Precision/Recall/F1 = 1.00), a further *12/65 (18.5%)* fell in 0.90–<1.00, and *5/65 (7.7%)* in 0.80–<0.90; only *2/65 (3.1%)* were in 0.30–<0.40, *1/65 (1.5%)* in 0–<0.10, and *1/65 (1.5%)* in chance–<0.60, yielding just *4.6%* at or below chance and *81.5%* at ≥ 0.90 accuracy. Macro-averaged across subjects (Top-1 per subject), test accuracy was 0.924 ± 0.183 with test F1 0.946 ± 0.154; macro-precision was 0.985 ± 0.124. These results indicate high patient-level separability with a small tail of difficult cases, suggesting that Hopfield networks have potential in differentiating AD and CN. Subjects (especially, Sub-43, 56, 59 < chance) resulted in incorrect prediction at large, questioning the robustness and generalizability of the trained AI models. Also, the accuracy bins show per-subject potential, not a deployable policy.

Besides Table 1, we also show results for two subjects (once from each class). While Figure 2 shows the epochwise training and testing performance metrics for AD/CN recognition (chance = 0.50), Table 2 shows the tabulated statistics for the top-5 performing epochs. These results are shown, as an example, for *Sub-001* (AD) and *Sub-040* (CN) under LOSO validation.

**TABLE 2.**
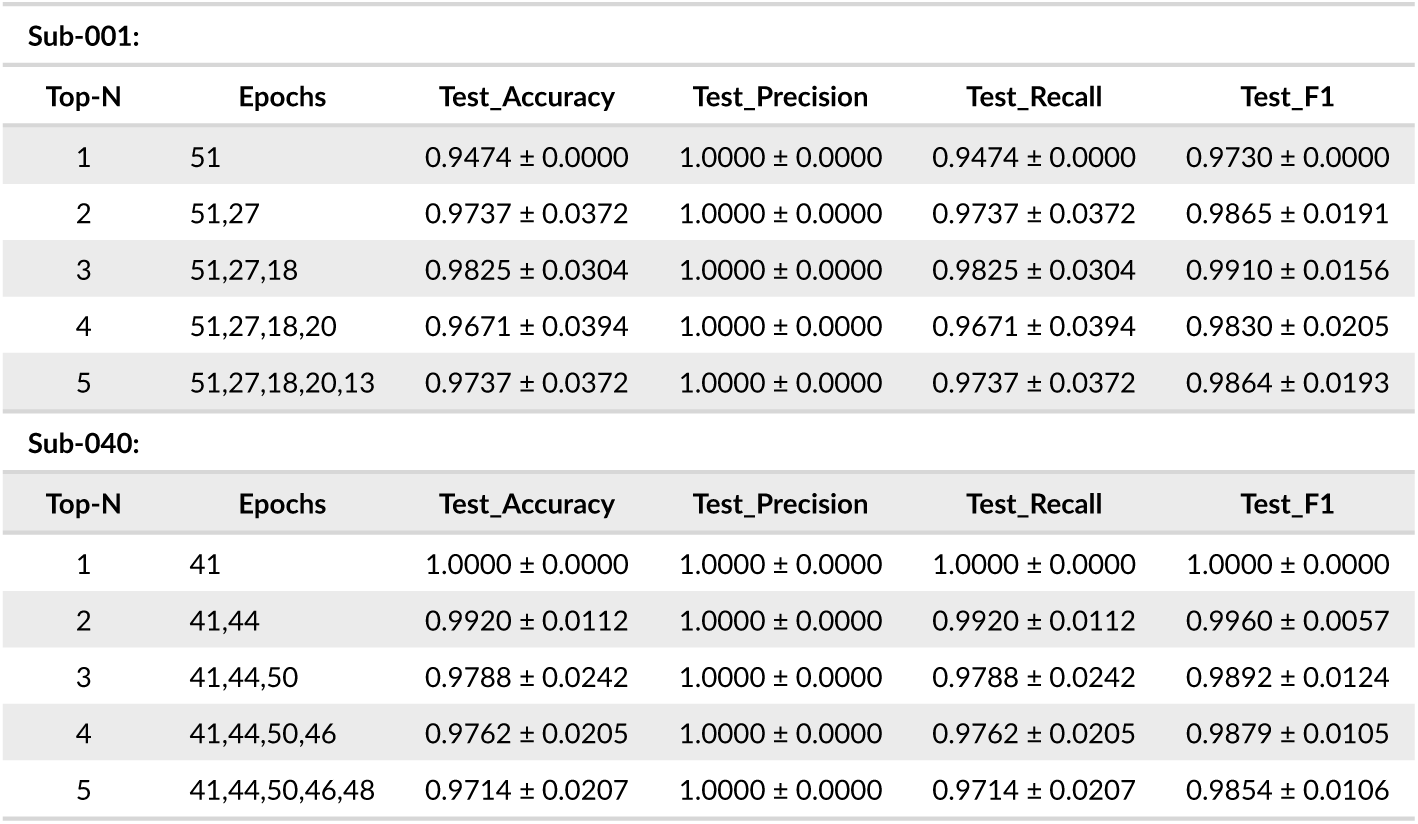
AD/CN: : Performance metrics up to Top-5 epochs with mean±std for Sub-001 and Sub-040.

**FIGURE 2.**
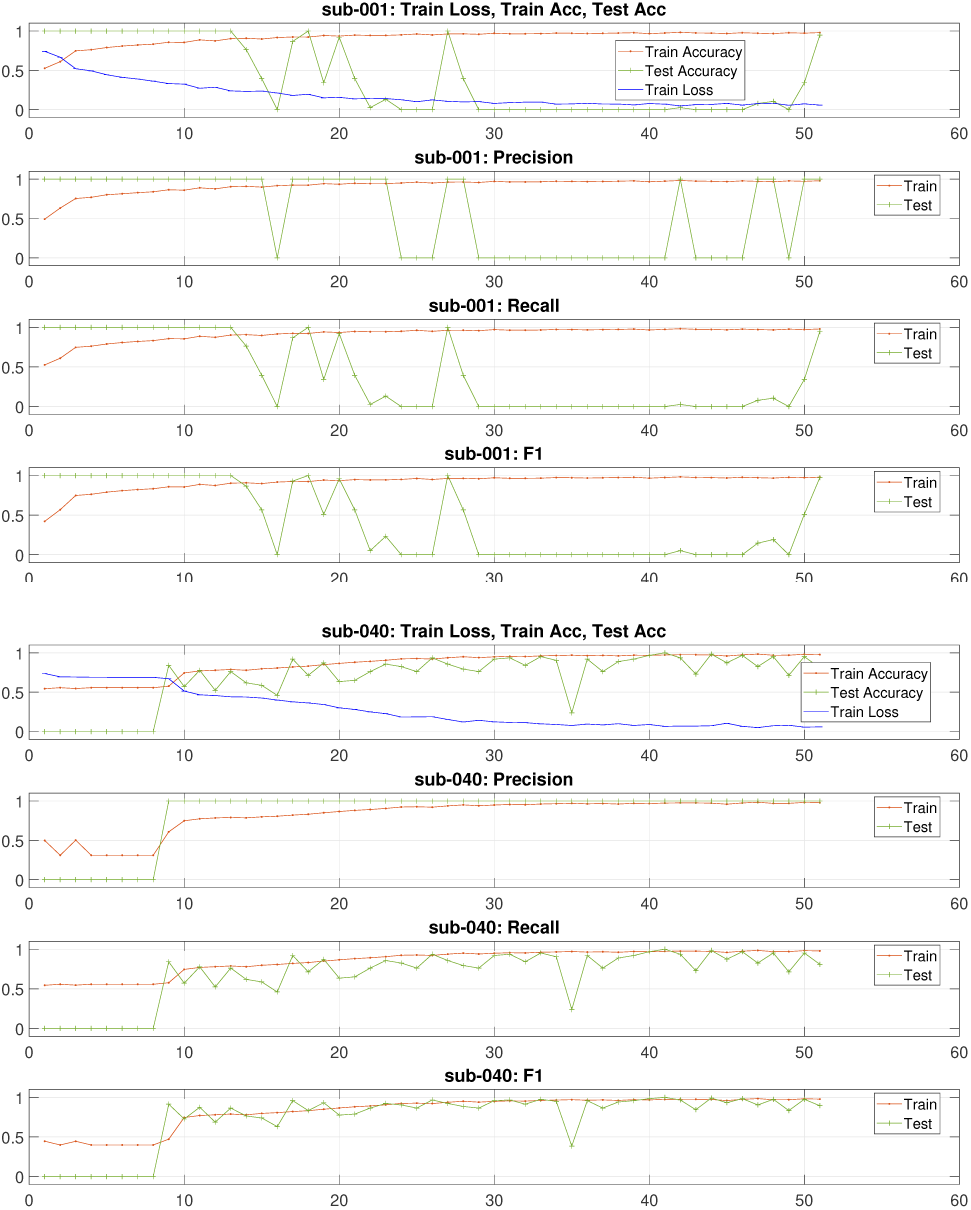
AD/CN-Training epochs-wise performance metrics for the training and testing for two subjects (sub-001, sub-040) in testing under LOSO validation.

We show all 51 epochs (Figure 2) for the two mentioned subjects in testing. For *Sub-001* (AD), performance was strikingly volatile: many early epochs were perfect (F1 = 1.000 at epochs 1–13, 18, 27), yet a prolonged mid-training collapse (approximately epochs 29–41) had near-zero accuracy and recall despite continued loss reduction. The **Top-1** checkpoint (epoch 51) yielded **Accuracy 0.9474, Precision 1.0000, Recall 0.9474, F1 0.9730**. Small checkpoint ensembles improved robustness for this subject: **Top-2** (epochs 51,27) Acc 0.9737 ± 0.0372, F1 0.9865 ± 0.0191; **Top-3** (51,27,18) Acc 0.9825 ± 0.0304, F1 0.9910 ± 0.0156; adding more epochs slightly diluted performance (**Top-5**: Acc 0.9737 ± 0.0372, F1 0.9864 ± 0.0193). In contrast, *Sub-040* (CN) showed well-behaved learning with a broad late-training high-performance band and a clean peak at **epoch 41 (Top-1 Accuracy 1.0000, Precision 1.0000, Recall 1.0000, F1 1.0000)**; checkpoint averaging remained strong with low variability (**Top-2** Acc 0.9920 ± 0.0112, F1 0.9960 ± 0.0057; **Top-3** Acc 0.9788 ± 0.0242, F1 0.9892 ± 0.0124; **Top-5** Acc 0.9714 ± 0.0207, F1 0.9854 ± 0.0106). Together, these paired examples highlight substantial patient-level heterogeneity - instability and thresholding artifacts in AD (Sub-001) versus stable gains in CN (Sub-040)—and motivate reporting **Top-N** checkpoint averages alongside **Top-1** to understand the reliability of patient-specific generalization.

### 4.2 | FTD/CN

Table 3 shows a fine-grained analysis of AD/CN results under LOSO validation. It summarizes subject-level classification performance for the FTD versus CN task, using the top-performing epoch for each subject. The distribution of subjects across accuracy bins demonstrates significant heterogeneity in model generalizability.

**TABLE 3.** FTD/CN-Binning of top-1 epoch for each subject-Further investigating.

| Accuracy Bins | Number of Subjects/Bin | Percent of Subjects/Bin | Subject ID | Test_Accuracy | Test_Precision | Test_Recall | Test_F1 |
| --- | --- | --- | --- | --- | --- | --- | --- |
| 0% -<10% | 3 | 5.7692 | 72,80,82 | 0.0167±0.0289 | 0.3333±0.5774 | 0.0167±0.0289 | 0.0317±0.0550 |
| 10% -<20% | 1 | 1.9231 | 79 | 0.1132±0.0000 | 1.0000±0.0000 | 0.1132±0.0000 | 0.2034±0.0000 |
| 20% -<30% | 0 | 0.0000 |  |  |  |  |  |
| 30% -<40% | 0 | 0.0000 |  |  |  |  |  |
| 40% -<chance | 2 | 3.8462 | 61,88 | 0.4565±0.0200 | 1.0000±0.0000 | 0.4565±0.0200 | 0.6266±0.0189 |
| chance -<60% | 2 | 3.8462 | 68,71 | 0.5723±0.0315 | 1.0000±0.0000 | 0.5723±0.0315 | 0.7278±0.0255 |
| 60% -<70% | 1 | 1.9231 | 69 | 0.6341±0.0000 | 1.0000±0.0000 | 0.6341±0.0000 | 0.7761±0.0000 |
| 70% -<80% | 1 | 1.9231 | 38 | 0.7069±0.0000 | 1.0000±0.0000 | 0.7069±0.0000 | 0.8283±0.0000 |
| 80% -<90% | 2 | 3.8462 | 39,43 | 0.8624±0.0112 | 1.0000±0.0000 | 0.8624±0.0112 | 0.9262±0.0064 |
| 90% -<100% | 22 | 42.3077 | 37,42,44,45,47,50,52,<br>53,54,55,56,57,59,62,<br>63,66,67,77,78,85,86,87 | 0.9423±0.0295 | 1.0000±0.0000 | 0.9423±0.0295 | 0.9701±0.0156 |
| =100% | 18 | 34.6154 | 40,41,46,48,49,51,58,60,64,<br>65,70,73,74,75,76,81,83,84 | 1.0000±0.0000 | 1.0000±0.0000 | 1.0000±0.0000 | 1.0000±0.0000 |
| OVERALL | 52 | 100 | All | 0.8465±0.2719 | 0.9615±0.1942 | 0.8465±0.2719 | 0.8809±0.2528 |

At the lower end, a small subset of subjects (3/52, 5.8%) consistently achieved near-zero accuracy (<0.1), indicating systematic misclassification rather than random error. An additional four subjects (7.7%) performed below or only slightly above chance level (<0.6), suggesting that the model failed to capture discriminative features for these individuals. In contrast, a large proportion of subjects (22/52, 42.3%) achieved high accuracy in the range of 0.9–<1.0, and 18 subjects (34.6%) reached perfect classification (1.0 across all metrics). This skewed distribution suggests that while the model is capable of near-perfect discrimination in many cases, performance collapses entirely for a nonnegligible minority of subjects. The overall mean accuracy was 0.8465 (±0.27), with balanced precision (0.9615), recall (0.8465), and F1-score (0.8809). These findings highlight the necessity of reporting subject-level variability, as group-level averages obscure clinically critical heterogeneity. For translation to practice, strategies to improve robustness in low-performing patients will be essential, since failure in even a small subset may have significant clinical consequences.

Besides Table 3, we also show results for two subjects (once from each class). While Figure 3 shows the epochwise training and testing performance metrics for AD/CN recognition (chance = 0.50), Table 4 shows the tabulated statistics for the top-5 performing epochs. These results are shown, as an example, for *Sub-039* (CN) and *Sub-073* (FTD) under LOSO validation.

**FIGURE 3.**
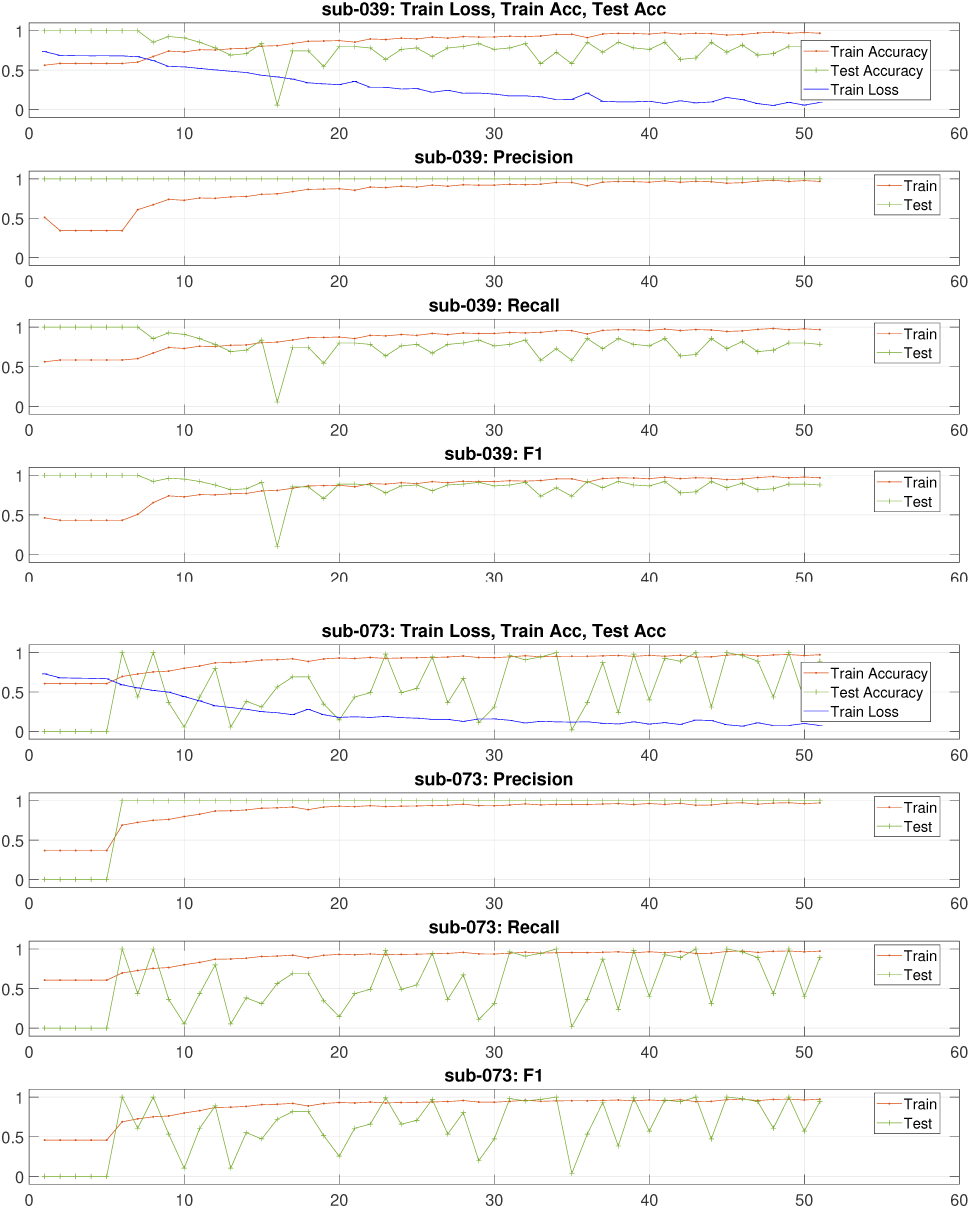
FTD/CN-Training epochs-wise performance metrics for the training and testing for two subjects (sub-039, sub-073) in testing under LOSO validation.

**TABLE 4.** FTD/CN: Performance metrics up to Top-5 epochs with mean±std for Sub-039 and Sub-073.

| Sub-039: |  |  |  |  |  |
| --- | --- | --- | --- | --- | --- |
| Top-N | Epochs | Test_Accuracy | Test_Precision | Test_Recall | Test_F1 |
| 1 | 41 | 0.8545 ± 0.0000 | 1.0000 ± 0.0000 | 0.8545 ± 0.0000 | 0.9216 ± 0.0000 |
| 2 | 41,38 | 0.8545 ± 0.0000 | 1.0000 ± 0.0000 | 0.8545 ± 0.0000 | 0.9216 ± 0.0000 |
| 3 | 41,38,44 | 0.8545 ± 0.0000 | 1.0000 ± 0.0000 | 0.8545 ± 0.0000 | 0.9216 ± 0.0000 |
| 4 | 41,38,44,50 | 0.8409 ± 0.0272 | 1.0000 ± 0.0000 | 0.8409 ± 0.0272 | 0.9134 ± 0.0163 |
| 5 | 41,38,44,50,49 | 0.8327 ± 0.0299 | 1.0000 ± 0.0000 | 0.8327 ± 0.0299 | 0.9085 ± 0.0179 |
| Sub-073: |  |  |  |  |  |
| Top-N | Epochs | Test_Accuracy | Test_Precision | Test_Recall | Test_F1 |
| 1 | 49 | 1.0000 ± 0.0000 | 1.0000 ± 0.0000 | 1.0000 ± 0.0000 | 1.0000 ± 0.0000 |
| 2 | 49,45 | 1.0000 ± 0.0000 | 1.0000 ± 0.0000 | 1.0000 ± 0.0000 | 1.0000 ± 0.0000 |
| 3 | 49,45,46 | 0.9879 ± 0.0210 | 1.0000 ± 0.0000 | 0.9879 ± 0.0210 | 0.9938 ± 0.0107 |
| 4 | 49,45,46,34 | 0.9909 ± 0.0182 | 1.0000 ± 0.0000 | 0.9909 ± 0.0182 | 0.9954 ± 0.0092 |
| 5 | 49,45,46,34,39 | 0.9891 ± 0.0163 | 1.0000 ± 0.0000 | 0.9891 ± 0.0163 | 0.9945 ± 0.0083 |

Across the two subjects, epoch-wise behavior shows structural metric effects and training instability: with single-class LOSO folds, accuracy = recall, precision is often 1.0 (few or no false positives), and F1 tracks recall. Sub-039 exhibits early perfect scores at relatively high loss (epochs 1–7; loss 0.73→0.67), then settles into a realistic band with accuracy/recall typically 0.55–0.85, including a collapse to 0.0545 at epoch 16; its Top-1 checkpoint (epoch 41) yields 0.8545 accuracy and 0.9216 F1, while Top-5 drops to 0.8327/0.9085, showing that simple checkpoint averaging does not stabilize performance. sub-073 is alternating between complete failure (epochs 1–5; accuracy 0.0) and perfect or near-perfect epochs (e.g., 6, 8, 31, 34, 39, 43, 45, 49), despite steadily decreasing loss; its Top-1 (epoch 49) is 1.0 across metrics and Top-3 remains near-ceiling (0.9879 accuracy, 0.9938 F1), implying high separability when the model lands in a favorable parameter region but poor reproducibility across epochs.

The epoch-wise volatility observed across these two subjects—including transitions from complete failure to perfect scores within the same individual—implies that a patient’s outcome can change meaningfully with checkpoint or run, which is unacceptable for clinical use.

### 4.3 | AD/FTD

Table 5 shows a fine-grained analysis of AD/FTD results under LOSO validation. It summarizes subject-level classification performance for the AD versus FTD task, using the top-performing epoch for each subject.

**TABLE 5.** AD/FTD-Binning of top-1 epoch for each subject-Further investigating.

| Accuracy<br>Bins | Number of<br>Subjects/Bin | Percent of<br>Subjects/Bin | Subject ID | Test_Accuracy | Test_Precision | Test_Recall | Test_F1 |
| --- | --- | --- | --- | --- | --- | --- | --- |
| 0%~<10% | 15 | 25.4237 | 67,69,71,72,73,75,77,79,80,82,83,84,85,86,87 | 0.0088±0.0217 | 0.2000±0.4140 | 0.0088±0.0217 | 0.0167±0.0405 |
| 10%~<20% | 4 | 6.7797 | 68,78,81,88 | 0.1401±0.0348 | 1.0000±0.0000 | 0.1401±0.0348 | 0.2446±0.0525 |
| 20%~<30% | 1 | 1.6949 | 70 | 0.2667±0.0000 | 1.0000±0.0000 | 0.2667±0.0000 | 0.4211±0.0000 |
| 30%~<40% | 0 | 0.0000 | - | - | - | - | - |
| 40%~<chance | 1 | 1.6949 | 76 | 0.4151±0.0000 | 1.0000±0.0000 | 0.4151±0.0000 | 0.5867±0.0000 |
| chance~<60% | 0 | 0.0000 | - | - | - | - | - |
| 60%~<70% | 0 | 0.0000 | - | - | - | - | - |
| 70%~<80% | 0 | 0.0000 | - | - | - | - | - |
| 80%~<90% | 1 | 1.6949 | 66 | 0.8571±0.0000 | 1.0000±0.0000 | 0.8571±0.0000 | 0.9231±0.0000 |
| 90%~<100% | 3 | 5.0847 | 8,10,74 | 0.9729±0.0139 | 1.0000±0.0000 | 0.9729±0.0139 | 0.9862±0.0071 |
| =100% | 34 | 57.6271 | 1,2,3,4,5,6,7,9,11,12,13,14,15,16,17,18,19, 20,<br>21,22,23,24,25,26,27,28,29,30,31,32,33,34,35,36 | 1.0000±0.0000 | 1.0000±0.0000 | 1.0000±0.0000 | 1.0000±0.0000 |
| OVERALL | 59 | 100 | All | 0.6636±0.4535 | 0.7966±0.4060 | 0.6636±0.4535 | 0.6800±0.4412 |

The distribution is strongly *skewed*, not unimodal. Out of 59 subjects, 34/59 (57.6%) achieve 100% accuracy at their Top-1 epoch, while 15/59 (25.4%) fall in the 0– < 10% bin and another 5/59 (8.5%) lie between 10% and 50%; only 1/59 sits just below chance (41.5%). There are *no* subjects between 50% and 80%, only 1/59 in 80– < 90%, and 3/59 in 90– < 100%. In practical terms, 37/59 (62.7%) reach ≥ 90% accuracy at their best epoch, while 20/59 (33.9%) are at or below chance (≤ 50%), indicating confident but brittle behavior that is *not* uniformly reliable across patients.

Besides Table 5, we also show results for two subjects (once from each class). While Figure 4 shows the epochwise training and testing performance metrics for AD/FTD recognition (chance = 0.50), Table 6 shows the tabulated statistics for the top-5 performing epochs. These results are shown, as an example, for Sub-005 (AD) and Sub-074 (FTD) under LOSO validation.

**FIGURE 4.**
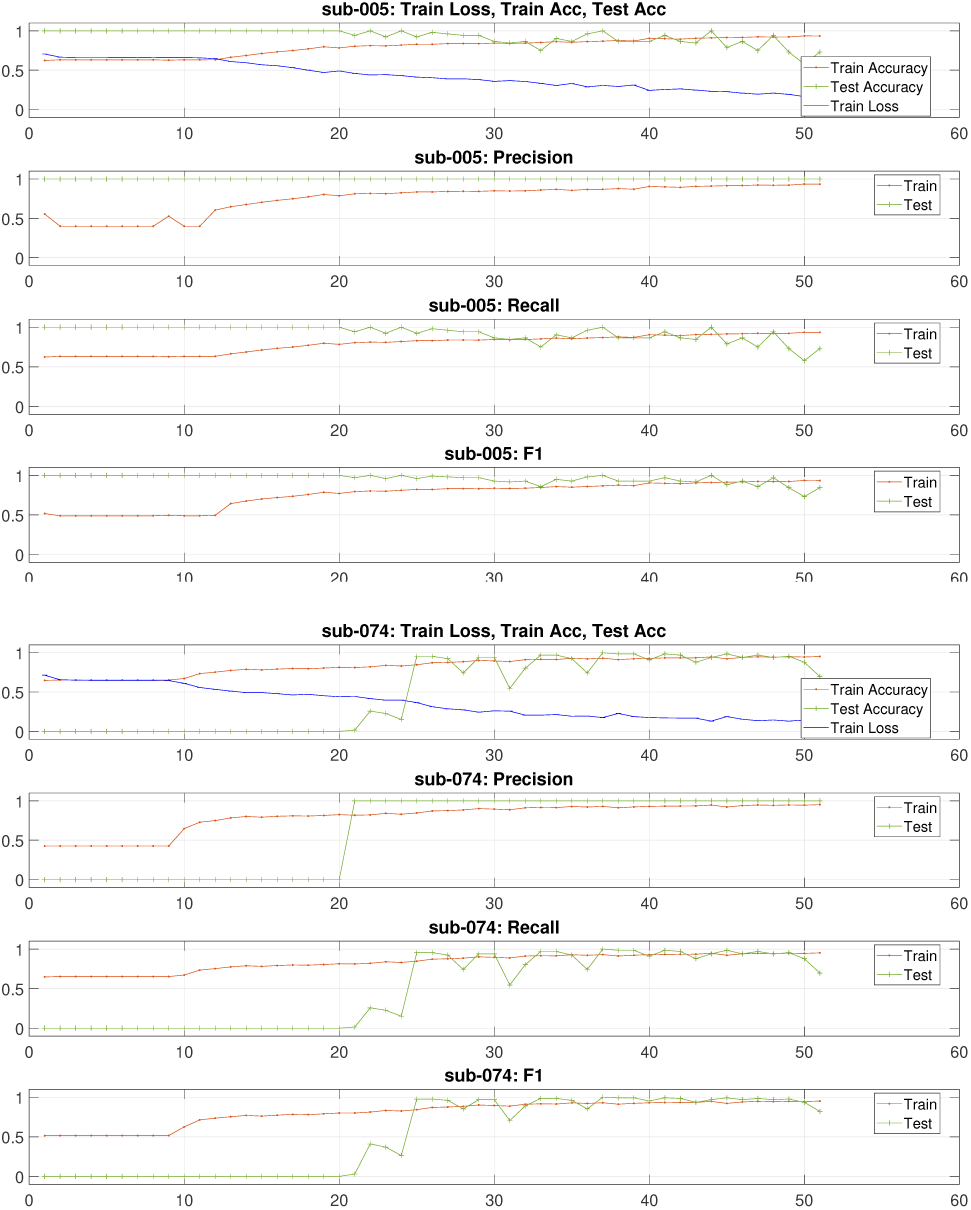
AD/FTD-Training epochs-wise performance metrics for the training and testing for two subjects (sub-005, sub-074) in testing under LOSO validation.

**TABLE 6.** AD/FTD: Performance metrics up Top-5 epochs with mean±std for Sub-005 and Sub-074.

| Sub-005: |  |  |  |  |  |
| --- | --- | --- | --- | --- | --- |
| Top-N | Epochs | Test_Accuracy | Test_Precision | Test_Recall | Test_F1 |
| 1 | 44 | 1.0000±0.0000 | 1.0000±0.0000 | 1.0000±0.0000 | 1.0000±0.0000 |
| 2 | 44,48 | 0.9711±0.0408 | 1.0000±0.0000 | 0.9711±0.0408 | 0.9851±0.0210 |
| 3 | 44,48,41 | 0.9615±0.0333 | 1.0000±0.0000 | 0.9615±0.0333 | 0.9802±0.0171 |
| 4 | 44,48,41,46 | 0.9375±0.0552 | 1.0000±0.0000 | 0.9375±0.0552 | 0.9671±0.0297 |
| 5 | 44,48,41,46,37 | 0.9500±0.0554 | 1.0000±0.0000 | 0.9500±0.0554 | 0.9737±0.0296 |
| Sub-074: |  |  |  |  |  |
| Top-N | Epochs | Test_Accuracy | Test_Precision | Test_Recall | Test_F1 |
| 1 | 47 | 0.9697±0.0000 | 1.0000±0.0000 | 0.9697±0.0000 | 0.9846±0.0000 |
| 2 | 47,49 | 0.9621±0.0107 | 1.0000±0.0000 | 0.9621±0.0107 | 0.9807±0.0056 |
| 3 | 47,49,37 | 0.9747±0.0232 | 1.0000±0.0000 | 0.9747±0.0232 | 0.9871±0.0118 |
| 4 | 47,49,37,44 | 0.9659±0.0259 | 1.0000±0.0000 | 0.9659±0.0259 | 0.9825±0.0133 |
| 5 | 47,49,37,44,41 | 0.9697±0.0240 | 1.0000±0.0000 | 0.9697±0.0240 | 0.9845±0.0123 |

The two shown subjects (Figure 4) display opposite learning trajectories with shared reliability concerns: *sub-005* attains perfect classification from the very first epoch and sustains it through epochs 1–20 (again at 24, 37, 44) while the training loss decreases (≈ 0.71 → 0.16); thereafter, test performance erodes, spending long intervals between 0.75 and 0.95 and collapsing late to 0.58–0.73 (epochs 50–51), consistent with boundary drift/overfitting to the training subjects despite continued optimization. *Sub-074* exhibits the mirror pattern: complete failure for ∼20 epochs, followed by an abrupt transition near epoch 25 into a sustained high-accuracy regime (multiple epochs ≥ 0.95).

The Top-5 summary in Table 6 shows that Sub-005 has perfect Top-1 (epoch 44) but mean accuracy declines and variance rises as N grows (Top-5 0.9500 ± 0.0554)—evidence of unstable checkpoints/overfit.

Sub-074 maintains high, tight Top-N (Top-5 0.9697 ± 0.0240) after a late improvement—indicating stable separability and reliable checkpoint selection. Subjects behave very differently, and the timing of learning matters: early 100% scores (sub-005) are not reliable for choosing a model, while late improvements (sub-074) mean we should let training run longer but pick the checkpoint using a separate, subject-independent validation set—not the test fold.

### 4.4 | AD/FTD/CN

We also experimented with three class settings, i.e., AD/FTD/CN. Table 7 shows a fine-grained analysis of AD/FTD/CN results under LOSO validation. It summarizes subject-level classification performance for AD versus FTD versus CN, using the top-performing epoch for each subject.

**TABLE 7.**
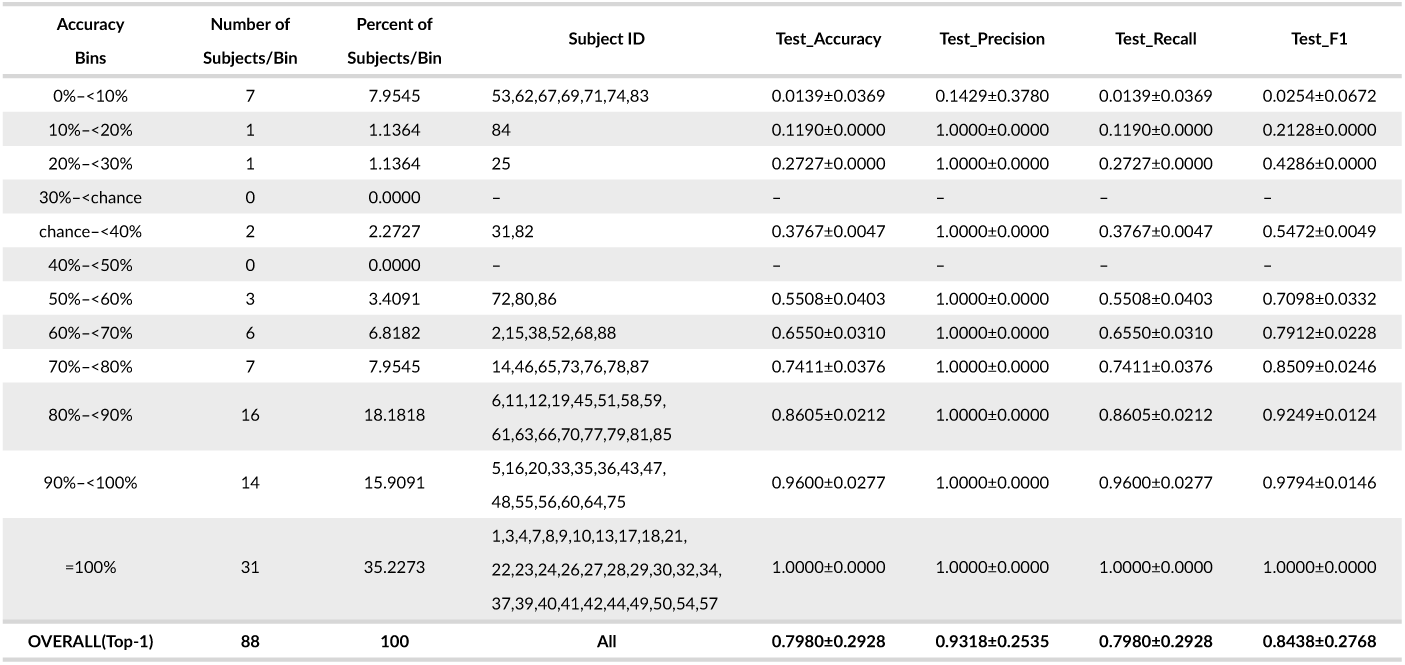
AD/FTD/CN-Binning of top-1 epoch for each subject-Further investigating.

| Accuracy<br>Bins | Number of<br>Subjects/Bin | Percent of<br>Subjects/Bin | Subject ID | Test_Accuracy | Test_Precision | Test_Recall | Test_F1 |
| --- | --- | --- | --- | --- | --- | --- | --- |
| 0%~<10% | 7 | 7.9545 | 53,62,67,69,71,74,83 | 0.0139±0.0369 | 0.1429±0.3780 | 0.0139±0.0369 | 0.0254±0.0672 |
| 10%~<20% | 1 | 1.1364 | 84 | 0.1190±0.0000 | 1.0000±0.0000 | 0.1190±0.0000 | 0.2128±0.0000 |
| 20%~<30% | 1 | 1.1364 | 25 | 0.2727±0.0000 | 1.0000±0.0000 | 0.2727±0.0000 | 0.4286±0.0000 |
| 30%~<chance | 0 | 0.0000 | – | – | – | – | – |
| chance~<40% | 2 | 2.2727 | 31,82 | 0.3767±0.0047 | 1.0000±0.0000 | 0.3767±0.0047 | 0.5472±0.0049 |
| 40%~<50% | 0 | 0.0000 | – | – | – | – | – |
| 50%~<60% | 3 | 3.4091 | 72,80,86 | 0.5508±0.0403 | 1.0000±0.0000 | 0.5508±0.0403 | 0.7098±0.0332 |
| 60%~<70% | 6 | 6.8182 | 2,15,38,52,68,88 | 0.6550±0.0310 | 1.0000±0.0000 | 0.6550±0.0310 | 0.7912±0.0228 |
| 70%~<80% | 7 | 7.9545 | 14,46,65,73,76,78,87 | 0.7411±0.0376 | 1.0000±0.0000 | 0.7411±0.0376 | 0.8509±0.0246 |
| 80%~<90% | 16 | 18.1818 | 6,11,12,19,45,51,58,59,<br>61,63,66,70,77,79,81,85 | 0.8605±0.0212 | 1.0000±0.0000 | 0.8605±0.0212 | 0.9249±0.0124 |
| 90%~<100% | 14 | 15.9091 | 5,16,20,33,35,36,43,47,<br>48,55,56,60,64,75 | 0.9600±0.0277 | 1.0000±0.0000 | 0.9600±0.0277 | 0.9794±0.0146 |
| =100% | 31 | 35.2273 | 1,3,4,7,8,9,10,13,17,18,21,<br>22,23,24,26,27,28,29,30,32,34,<br>37,39,40,41,42,44,49,50,54,57 | 1.0000±0.0000 | 1.0000±0.0000 | 1.0000±0.0000 | 1.0000±0.0000 |
| OVERALL(Top-1) | 88 | 100 | All | 0.7980±0.2928 | 0.9318±0.2535 | 0.7980±0.2928 | 0.8438±0.2768 |

The distribution of subjects in different accuracy bins is highly skewed: 31/88 (35.2%) achieve 100% accuracy; 45/88 (51.1%) are ≥ 90%; and 61/88 (69.3%) are ≥ 80%. At the lower end, 11/88 (12.5%) are ≤ 40%, including 7/88 (8.0%) in the 0– < 10% bin. The 30%– < chance and 40%– < 50% bins are empty, suggesting a valley between easy and hard subjects rather than a smooth continuum. Per-bin *F*_1_ increases with accuracy, while precision is saturated near 1.0 in most bins and thus offers limited discriminative value for difficulty stratification. The overall Top-1 mean (±SD) is 0.798 ± 0.293. These patterns indicate inter-subject heterogeneity and potential susceptibility to subject-specific artifacts or split idiosyncrasies; for reporting and selection, prefer subject-level confidence intervals, targeted audits of the hard tail (signal quality, subject-level priors, leakage checks, non-overlapping windows), and a test-agnostic, validation-driven checkpoint policy to avoid overfitting to abundant “easy” cases.

Besides Table 7, we also show results for three subjects (once from each class). While Figure 5 shows the epochwise training and testing performance metrics for AD/FTD/CN recognition (chance = 0.33), Table 8 shows the tabulated statistics for the top-5 performing epochs. These results are shown, as an example, for Sub-011 (AD), Sub-041 (CN), and Sub-081 (FTD) under LOSO validation.

**FIGURE 5.**
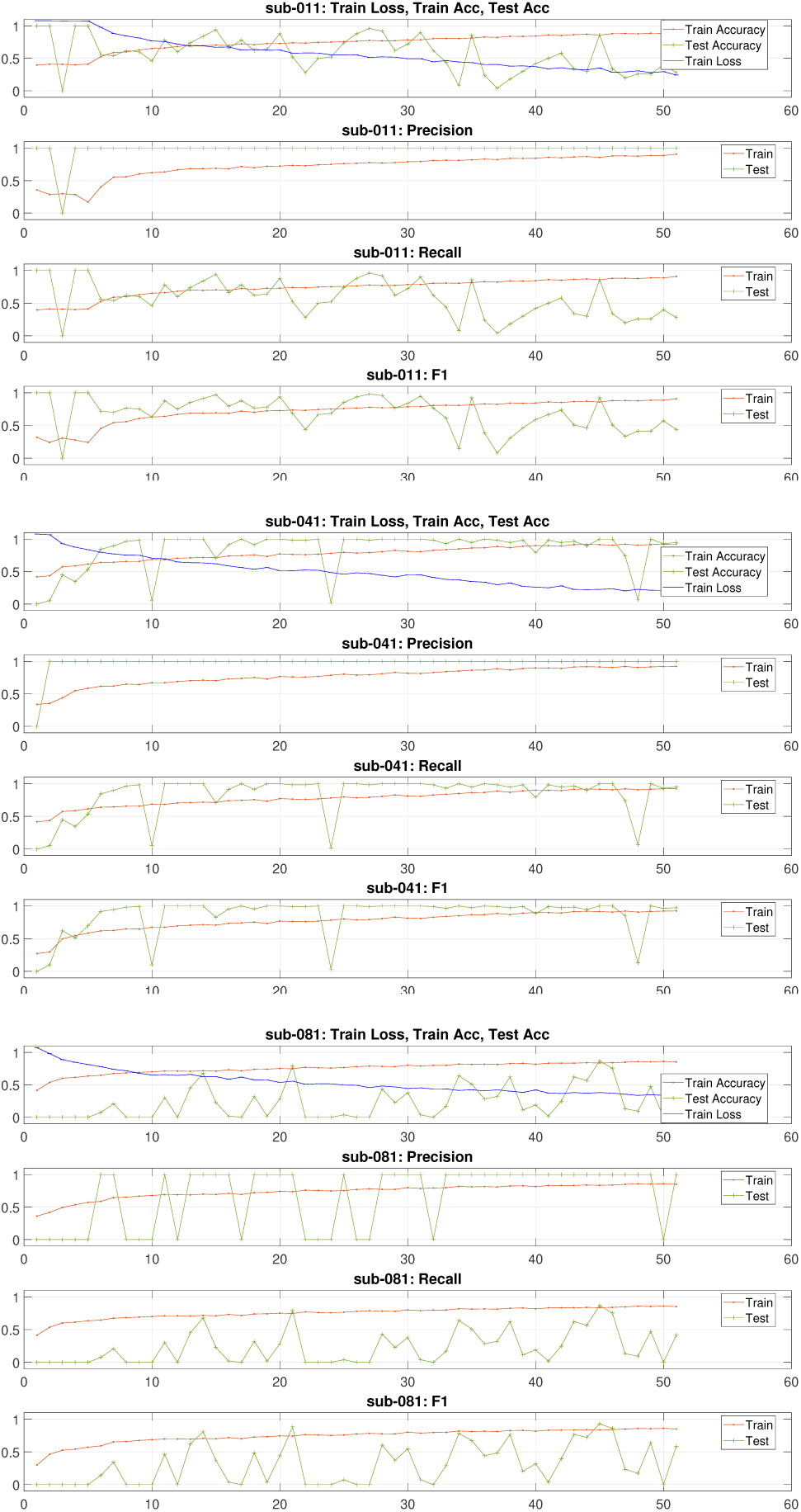
AD/FTD/CN-Training epochs-wise performance metrics for the training and testing for three subjects (sub-011, sub-041, sub-088) in testing under LOSO validation.

**TABLE 8.**
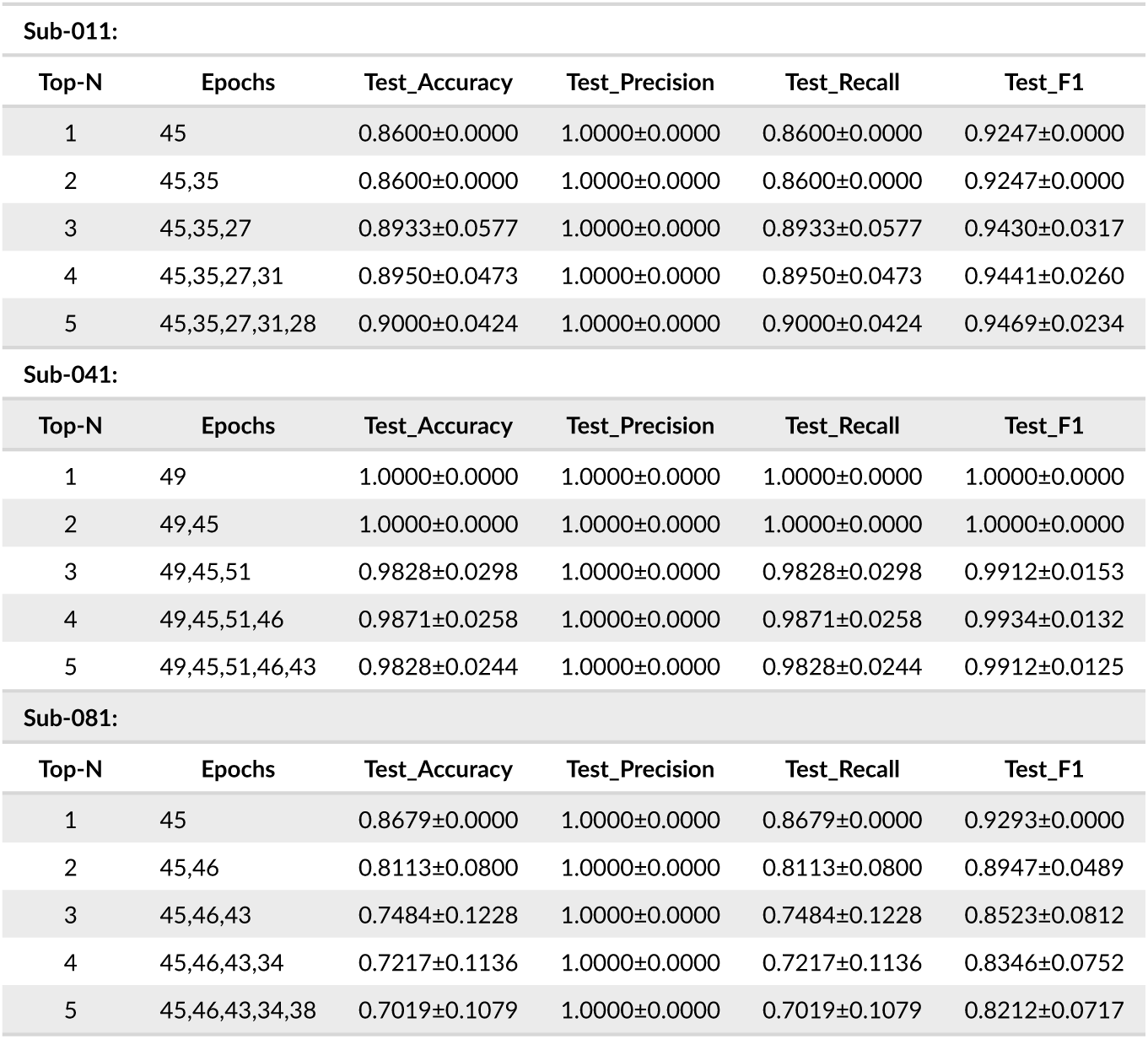
AD/FTD/CN: Performance metrics up to Top-5 epochs with mean±std for Sub-011, Sub-041, and Sub-088.

It can be noticed from Figure 5 that a decoupling between decreasing train loss and jagged test accuracy indicates variance-dominated generalization and unstable decision boundaries rather than smooth learning.

Across the three subjects, Top-N behavior diverges sharply: Sub-011 shows a narrow plateau of strong check-points where small-N ensembling is beneficial (Top-1 0.86 → 0.90 at Top-5), Sub-041 is an “easy” fold dominated by a single excellent checkpoint where adding weaker epochs dilutes performance (1.00 → 0.9828), and Sub-081 is a high-variance fold with sparse good epochs where ensembling is harmful (0.8679 → 0.7019).

A cross-fold consensus around epoch 45 (in all Top-2 sets; optimal for sub-081) makes a pre-defined global checkpoint at 45 a reasonable candidate, but the observed volatility (occasional collapses despite low train loss) argues for a validation-driven, subject-independent selection policy and for emphasizing subject-level stability over large-N averages.

## 5 | STRUCTURED LOGS

To ensure transparency, reproducibility, and clinical interpretability of our results, we maintain structured training and evaluation logs for all experiments. These logs serve two main purposes:

1. They allow independent verification that each model was trained and tested without data leakage between subjects, and
2. They provide detailed, per-subject performance trends across all training epochs, enabling clinicians and researchers to assess whether the trained models are robust and generalizable.

These logs are provided in two formats: (i) a standard Excel workbook (.xlsx) and (ii) a macro-enabled Excel workbook (.xlsm) with interactive features; Figure 6 (a)-(e) shows the screenshots of the sheets for different subjects.

**FIGURE 6.**
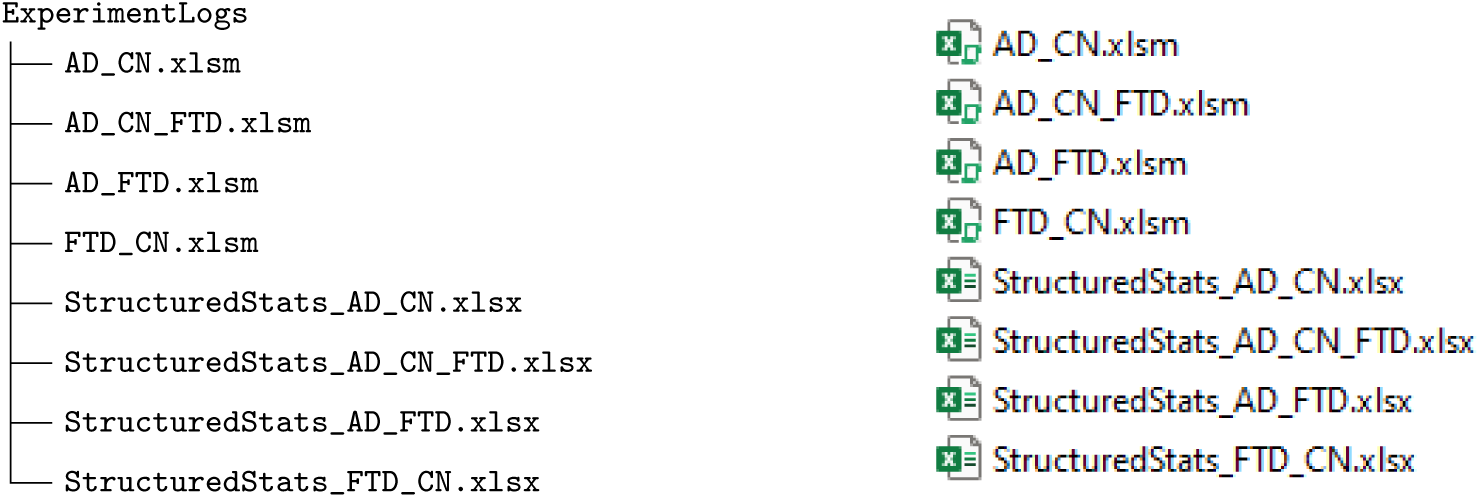
Folder structure showing macro-enabled logs (.xlsm) and corresponding structured statistics files (.xlsx). Left: vector representation. Right: actual folder screenshot for reference.

1. **Excel workbook (*.xlsx)**
  - Contains a single sheet named `StructuredStats`.
  - Presents a clean table with one row per training epoch, including:
    – Epoch, Subject (held-out subject ID),
    – Training metrics: loss, accuracy, precision, recall, F1-score,
    – Testing metrics: accuracy, precision, recall, F1-score.
  - The table allows clinicians to see how model performance changes over the 51 training epochs for each test subject.
2. **Macro-enabled workbook (*.xlsm)**: It contains multiple sheets for a deeper inspection and visualization. An example screenshot is also shown in Figure 7.
  - **RawLogs** – Full record of the training process as printed from the analysis scripts, including the list of training subjects, the single test subject, per-epoch metrics, and confusion matrices. This sheet demonstrates that no subject overlap occurs between training and testing.
  - **validLogs** – A filtered version of RawLogs containing only the essential per-epoch metrics for the training and test subject, making it easier to scan without auxiliary messages.
  - **StructuredStats** – Same table as in the .xlsx file, included here for reference and linkage to other sheets.
  - **SubjectSpecificStatsAndPlots** – Interactive view where entering a subject ID in the designated cell automatically fetches that subject’s full training and testing metrics from StructuredStats. It also generates plots of accuracy, precision, recall, and F1-score across epochs for both training and testing for extended learning and deeper analysis.
    – This sheet includes two extra columns:
      * Epoch_Score_weighted – a combined score reflecting both training and testing performance while penalizing high training loss. Higher scores indicate better generalization, with -1 being the poorest.
      * Ranking – the position of each epoch based on Epoch_Score_weighted (best ranking = 1).
    – Navigation tip: Changing a subject ID in this sheet (in the cell next to *Fetch Subject from StructuredStats*) immediately updates the plots and ranking table for that subject.
    – Top-N statistics:

* For N=1,2,3,4,5, the top N ranked epochs are selected (excluding invalid scores). Provided on the same sheet; please scroll down.
* For each N, the mean ± standard deviation is calculated for all metrics.
* The results are displayed in a compact table, allowing the reader to see not just the single best epoch but also the stability and variability of the next-best epochs.
* Clinically, these statistics help assess whether the high performance of a model is consistent across multiple training checkpoints, which is crucial for reliability in deployment.
3. **How to navigate the logs**
  - Use the .xlsx file for a quick overview of numerical results.
  - Use the .xlsm file for interactive exploration:
    1. Start in `StructuredStats` to get the complete table of results.
    2. Move to `SubjectSpecificStatsAndPlots` to investigate a single subject in detail and review top-performing epochs.
    3. Refer to `validLogs` or `RawLogs` if you need to confirm the original training process and subject allocation.

**FIGURE 7.**
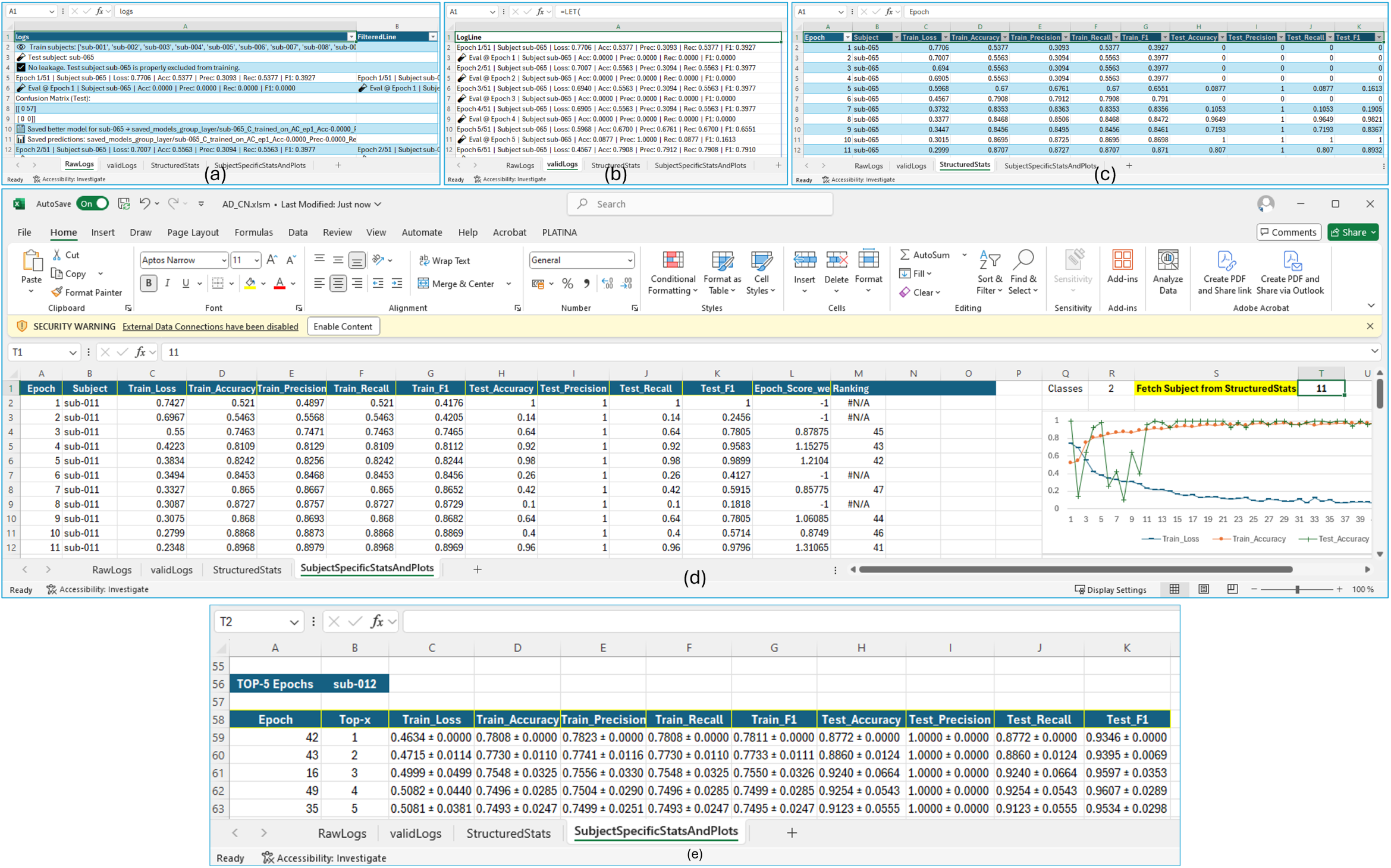
The screenshots of logs: (a) *RawLogs*, (b) *validLogs*, (c) *StructuredStats*, (d) *SubjectSpecificStatsAndPlots*, (e) *Top-5 ranked epochs with mean and standard deviation*.

These structured logs are intended to make the model’s development process fully auditable, ensuring that both technical reviewers and clinical experts can verify that performance claims are grounded in transparent and traceable evidence.

## 6 | DISCUSSION

This study examined whether high group-level performance of EEG-based deep learning (DL) models translates into dependable, per-patient behavior suitable for clinical decision-making. Across four diagnostic settings (AD/CN, FTD/CN, AD/FTD, and AD/FTD/CN), we observed substantial subject-level heterogeneity and checkpoint instability that are invisible in aggregate metrics yet critical for deployment. In brief, (i) some individuals were classified perfectly from the very first epoch onward, (ii) others remained persistently below chance, and (iii) many showed volatile trajectories where the predicted label for a given subject could change across epochs despite monotonically decreasing training loss. These patterns emerged under a rigorously enforced LOSO protocol with programmatic leakage checks, identical hyperparameters, and fixed training length, indicating that they are not artifacts of evaluation practice but reflect genuine brittleness in subject-level generalization.

### What the results reveal about subject-level generalizability

The AD/CN task displayed a strongly right-skewed Top-1 distribution (63.1% of subjects at 100% accuracy; 81.5% at ≥0.90), alongside a small but non-negligible tail at or below chance, demonstrating that “easy” subjects can dominate averages while a minority experiences systematic failure. A similar picture held for FTD/CN (34.6% perfect; 5.8% nearzero accuracy), and was most extreme in AD/FTD, where 57.6% of subjects achieved 100% yet 33.9% were at or below chance—an unmistakable bimodality rather than unimodal difficulty. In the three-class AD/FTD/CN setting, 35.2% of subjects reached 100%, but 12.5% were ≤0.40, again indicating a valley between very separable and persistently hard cases. Taken together, these distributions show that strong means can coexist with clinically unacceptable variance: a tool that works “on average” may still fail reliably for specific individuals.

### Checkpoint choice and metric structure matter

Epoch-wise plots for representative subjects illustrate two reliability hazards. First, “early perfect” cases (e.g., perfect test scores across many early epochs) can later degrade as the boundary drifts toward training subjects, so Top-1 at a lucky epoch is not a robust selection rule. Second, some subjects transition abruptly from failure to near-perfect performance mid-training, confirming that a patient’s outcome may depend on the particular checkpoint rather than stable learning. Small Top-*N* checkpoint averages sometimes improve stability (e.g., AD/CN Sub-001 and FTD/CN Sub-073) but can also dilute performance (e.g., FTD/CN Sub-039; AD/FTD/CN Sub-081), reinforcing that ensembling within the *test* fold is an analysis device rather than a deployable policy. A validation-driven, subject-independent checkpoint criterion is therefore mandatory.

A further nuance is metric structure under LOSO. Because each test fold contains windows from a single true class, accuracy equals recall, precision is often saturated at 1.0 (few false positives by construction), and F1 largely tracks recall. Precision, therefore, has limited discriminative value for difficulty stratification in this setting; reporting accuracy/recall (and confidence intervals) is more informative.

### Clinical implications

For clinical translation, reliability at the patient level is the currency of safety. Our findings indicate that (i) per-subject reporting is essential, (ii) any deployment must include an uncertainty-aware operating mode (e.g., calibrated confidence with abstention/referral when uncertainty is high), and (iii) validation policies must be independent of the test fold. Subject-level binning is useful to expose heterogeneity but “shows per-subject potential, not a deployable policy,” and primary claims should rely on selection rules that do not condition on test outcomes (e.g., fixed or validation-driven checkpointing).

### Why does heterogeneity arise?

We verified strict subject separation, reproducible training, and consistent hyperparameters, suggesting that failures are not due to leakage or procedural variance. Instead, heterogeneity likely reflects (a) idiosyncratic EEG characteristics (signal quality, physiology, state), (b) class-conditional confounds that create spuriously separable “easy” cases, and (c) instability of the learned boundary under limited data and class imbalance. The Hopfield-based architecture captures useful patterns (as evidenced by the large “easy” mass), but the tails reveal sensitivity to subject-specific artifacts and optimization idiosyncrasies.

### Strengths

Key strengths include a strict LOSO evaluation aligned to the clinical use case (generalization to unseen patients), explicit programmatic safeguards against leakage, and comprehensive, auditable logs that expose epoch-wise behavior and enable independent verification. We also release structured per-subject logs and code to facilitate reproducibility and critical reanalysis.

### Limitations

Our analyses use a single-center, resting-state dataset with 88 subjects; external validation across centers, hardware, and acquisition protocols is needed to assess transportability. We trained for a fixed 51 epochs with no hyperparameter tuning or early stopping; while this choice avoided overfitting to a validation subset and made instability more salient, it may understate the best attainable stability with stronger regularization and tuning. Finally, Top-*N* summaries in this paper are used diagnostically; any real-world model selection must be validation-driven and test-agnostic.

### Recommendations for robustification and reporting

Based on the above, we recommend that future EEG-AI studies targeting clinical use:

1. Adopt subject-independent checkpointing (fixed epoch or validation fold) and report per-subject confidence intervals alongside group means.
2. Include an uncertainty-aware operating point with abstention/referral for low-confidence patients; audit the hard tail with targeted quality checks (signal quality, channel-wise artifacts, non-overlapping window checks).
3. Prefer metrics that reflect LOSO structure (accuracy/recall and F1), and avoid overinterpreting precision when it is structurally saturated.
4. Explore robustness strategies that act on subject variability: subject-balanced sampling, mixup/cutout across subjects, distributionally robust optimization, channel dropout and spectral-shift augmentations, and calibration with leakage-safe normalization.
5. Perform multi-center external validation and prospective testing to quantify transport and drift.
6. Continue to use and release structured, auditable logs so that epoch-wise instability and per-subject outcomes can be independently scrutinized.

### Conclusion

Hopfield-enhanced DL models are a good candidate to model EEG signals for Alzheimer’s and dementia. It can achieve impressive average performance, but averages conceal clinically important heterogeneity and checkpoint sensitivity. Patient-level validation, test-agnostic selection, uncertainty-aware operation, and transparent reporting are not optional extras—they are prerequisites for safe translation of EEG-AI into dementia care.

### Future directions

We will test the approach across multiple EEG datasets if results hold in newer ones and over time. We will also explore new “foundational EEG models”—large, general models trained on many EEG recordings—as a stronger starting point, so less new data is needed and the system adapts more easily to different devices and patient groups. We will compare these models with our current approach and check that their results line up with routine clinical measures such as memory tests and standard brain exams. Finally, we plan to share clear documentation, code, and subject-level summaries so other teams can repeat and improve this work.

## Data Availability

https://openneuro.org/datasets/ds004504/versions/1.0.7
Andreas Miltiadous, Katerina D. Tzimourta, Theodora Afrantou, Panagiotis Ioannidis, Nikolaos Grigoriadis, Dimitrios G. Tsalikakis, Pantelis Angelidis, Markos G. Tsipouras, Evripidis Glavas, Nikolaos Giannakeas, and Alexandros T. Tzallas (2024). A dataset of EEG recordings from: Alzheimer's disease, Frontotemporal dementia and Healthy subjects. OpenNeuro. [Dataset] doi: doi:10.18112/openneuro.ds004504.v1.0.8

## Abbreviations

EEG: Electroencephalography
LOSO: Leave-one-subject-out
AD: Alzheimer’s
CN: Healthy controls
FTD: Frontotemporal dementia
AI: Artificial Intelligence
ML: Machine Learning.

## ACKNOWLEDGMENTS

The study was funded by Stiftelsen Promobilia under NeuraMind project with grant number A23110.

## AUTHOR CONTRIBUTIONS

All authors listed herein have made a substantial, direct, and intellectual contribution to the work, and all approve publication. Rajkumar Saini: Project administration; conceptualization; methodology; experimentation; data analysis, and writing-original draft. Vibha Gupta: Conceptualization, and review & editing. Foteini Liwicki: Project administration; supervision, and review & editing. Sumit Rakesh: AI-cluster administration, Technical lead, and review & editing. Debashis Das Chakladar: Review & editing. Hamam Mokayed: Review & editing. Sarthak Acharya: Review & editing. Daljit Singh: Review & editing. Araz Rawshani: Review & editing.

## DATA AND CODE AVAILABILITY

The dataset is publicly available at OpenNeuro data platform with dataset identifier ‘ds004504’. The experimental logs and implementation codes are provided as supplementary material to this article.

